# Leveraging Stacked Classifiers for Multi-task Executive Function in Schizophrenia Yields Diagnostic and Prognostic Insights

**DOI:** 10.1101/2024.12.05.24318587

**Authors:** Tongyi Zhang, Xin Zhao, B.T. Thomas Yeo, Xiaoning Huo, Simon B. Eickhoff, Ji Chen

## Abstract

Cognitive impairment is a central characteristic of schizophrenia. Executive functioning (EF) impairments are often seen in mental disorders, particularly schizophrenia, where they relate to adverse outcomes. As a heterogeneous construct, how specifically each dimension of EF to characterize the diagnostic and prognostic aspects of schizophrenia remains opaque. We used classification models with a stacking approach on systematically measured EFs to discriminate 195 patients with schizophrenia from healthy individuals. Baseline EF measurements were moreover employed to predict symptomatically remitted or non-remitted prognostic subgroups. EF feature importance was determined at the group-level and the ensuing individual importance scores were associated with four symptom dimensions. EF assessments of inhibitory control (interference and response inhibitions), followed by working memory, evidently predicted schizophrenia diagnosis (area under the curve [AUC]=0.87) and remission status (AUC=0.81). The models highlighted the importance of interference inhibition or working memory updating in accurately identifying individuals with schizophrenia or those in remission. These identified patients had high-level negative symptoms at baseline and those who remitted showed milder cognitive symptoms at follow-up, without differences in baseline EF or symptom severity compared to non-remitted patients. Our work indicates that impairments in specific EF dimensions in schizophrenia are differentially linked to individual symptom-load and prognostic outcomes. Thus, assessments and models based on EF may be a promising tool that can aid in the clinical evaluation of this disorder.

## Introduction

Schizophrenia is a serious mental health condition that can severely impair an individual’s functioning and quality of life. Individuals with schizophrenia present a wide range of symptoms of varying severity. These symptoms feature hallucinatory and delusional experiences termed “positive symptoms” as well as negative symptoms which manifest as atypical emotional and social behaviors. Distinct from negative symptoms, though related, cognitive symptoms involve the impairment of mental functions related to memory, attention, and executive tasks. This greatly affects the ability to live independently given the difficult to treat these symptoms using the currently available antipsychotic medications^1^. The cognitive symptoms in schizophrenia are a core aspect of psychopathology; they are considered a trait marker that emerges in the prodromal phase and persists throughout the illness^2^, unlike those manifested in affective psychotic disorders or drug-induced psychosis where cognitive deficits are epiphenomenal. However, cognitive performance in patients with schizophrenia is heterogeneous, and can vary from virtually unaffected to severely impaired^3,4^. While there is mixed evidence regarding the impairments in some cognitive domains, executive dysfunction is pervasively abnormal in schizophrenia. Previous studies have consistently indicated that mild-to-severe deficits in processes are related with executive functions (EFs)^5^.

EF represents a series of higher-order cognitive processes that involve impulse control and behavior orchestration^6^. Deficits in this domain can hinder goal-directed activity and contribute to aggression, violence, and poor compliance to medication in patients with schizophrenia, leading to worse clinical outcomes^7^. In the field of psychiatry, prognostic prediction remains a significant challenge in research and clinical practice, though it is crucial for early assessment and intervention. Previous studies employing baseline neuroimaging, genetic, or clinical data only approached chance-level accuracy in most cases^8,9^. This emphasized the lack of reliable markers for tracking the disease trajectory^10,11^. EF deficits emerge in the early stages (e.g., ultra-high risk, first episode) of schizophrenia^12,13^ and have been related to disease progression, symptom severity, and recovery of social and occupational skills^14^. Hence, they might be potential markers for tracking the clinical courses and prognostic statuses of schizophrenia.

A broad range of EF impairments has been associated with this disorder, including increased difficulties in inhibiting automatic responses and switching to new ones, reduced cognitive flexibility, and disturbances in the maintenance and updating of goal-related or rule-based information in working memory. These actually align well with the three-factor model of EF proposed by Miyake et al.^15^, which characterizes 1) interference inhibition and response inhibition; 2) cognitive flexibility and switching; and (3) working memory updating and maintenance. This three-dimensional representation of EF functions robustly captures individual variation in EF subcomponents across a broad spectrum of age groups and clinical cohorts, including patients with psychiatric disorders^16,17^. These dimensions of EF differ in concepts and neurobiological substrates, highlighting the need to consider and assess these dimensions, while studying their correlations with diagnostic and prognostic aspects of schizophrenia. Such finer characterization of EF functions may assist in understanding different psychopathological processes in schizophrenia (e.g., disorganization symptoms), which manifest as difficulties in the goal-directed sequencing of thoughts and behaviors^18,19^. These symptoms are linked to the cognitive dimension in our recently introduced four-dimensional representation (positive, negative, cognitive, and affective) of schizophrenia psychopathology, as generalizable across populations and clinical settings^20^. Disturbances in EF have likewise been implicated in negative and positive symptoms. Firstly, failure in effectively monitoring volitional behaviors and inhibiting false inference in predictive processing would have consequences on positive symptoms (e.g., hallucinations and delusions)^21,22^. Secondly, cognitive rigidity hampers adjustment of thoughts and actions for environmental volatility^23^. Thirdly, impaired working memory updating and maintenance has been linked to poor abstract thinking^24^, which can increase the risk of developing negative symptoms (e.g., apathy and diminished expressive behavior) commonly observed in schizophrenia^23,25^. However, some dimensions of EF may be more severely affected than others in schizophrenia^26^. Currently, it remains unclearly the abnormalities in which EF dimensions characterize schizophrenia and would play a role in providing prognostic information.

Assessment of the underlying processes of the dimensions of EF is a challenge. Currently available tools (e.g., Cambridge Neuropsychological Test Automated Battery^27^ and the National Institutes of Health Toolbox^28^) do not feature a comprehensive assessment to cover the various dimensions of EF. Furthermore, assessments included in available neuropsychological batteries (e.g., Delis–Kaplan Executive Function System) are paper tests rather than experiments. Consequently, they do not offer a trial-by-trial based dynamic quantification, e.g., the reaction time in a sequential task. Trial-by-trial responses help detect subtle cognitive impairments in schizophrenia, including EF^29,30^. In addition, the cognitive symptom items routinely used in clinical practice, including EF rating such as in the Positive and Negative Syndrome Scale (PANSS), are retrospective based on information collected from patient interviews or contributions by relatives. Comparatively, task paradigms drawn from the cognitive psychology literature provide objective trial-by-trial tests that facilitate measuring particular cognitive functions with likely improved sensitivity and specificity^16,31^. Such a tailored assessment strategy would be ideal for investigating the diagnostic and prognostic value of EF dimensions in schizophrenia by establishing classification models. Previous psychiatric machine learning studies mainly considered single algorithms, such as support vector machine (SVM) or random forest (RF), comparing their respective accuracies^32^. Methods such as stacking, a mainstay multi-view learning approach, may be another strategy for improving model performance^33^.

In this study, we systematically applied six well-established behavioral paradigms to assess individual baseline functions along the three EF dimensions (i.e., inhibitory control, working memory maintenance and updating, and cognitive flexibility) to determine their consistency in characterizing patients with schizophrenia and their prognostic statuses at follow-up. This is tested via establishing diagnostic and prognostic classification models using machine learning methods: SVM, RF, Adaptive Boosting (AdaBoost), and their stacked model with a stringent nested cross-validation (CV) and independent testing. The importance of EF feature contributions to classification models was determined via the SHapley Additive exPlanations (SHAP) approach, which facilitates the identification of important features at both the group- and individual-levels. This approach enables a link between feature importance and individual psychopathology (Fig. 1).

**Fig. 1.**
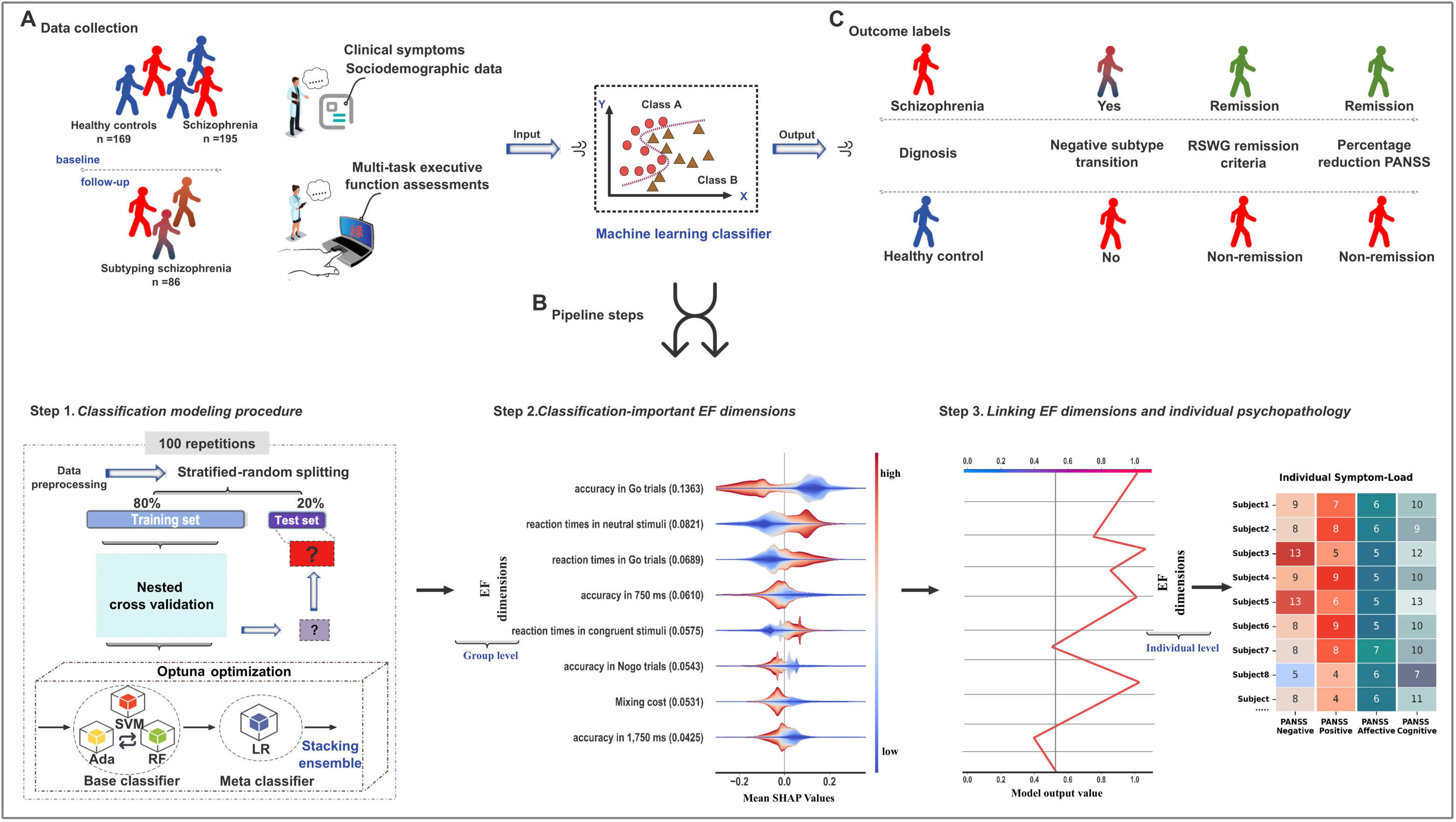
Study overview. **A).** The diagnostic model included the recruitment of patients with schizophrenia, further stratified into positive and negative symptom subtypes, and healthy controls. All participants underwent a comprehensive battery of tests and assessments tailored to their respective groups. The prognostic model was developed from a cohort of 86 patients with schizophrenia who completed a standard treatment regimen within 4–6 weeks of hospitalization. At follow-up, patients were evaluated using the RSWG criteria as the primary outcome measure. Additionally, three widely accepted definitions in the field were incorporated to comprehensively classify treatment response (i.e., 25%, 35%, and 50% symptom reduction thresholds), as well as changes in positive and negative symptom subtypes. **B).** Data underwent preprocessing, followed by a stratified random division into an 80% discovery dataset and a 20% test dataset, balanced for diagnostic and remission outcomes. The discovery set was subjected to nested cross-validation, with model performance assessed on the test set using various metrics. To reduce splitting variance, this procedure was repeated 100 times. **C).** SHAP values assigned to executive function features indicated their importance in model predictions, with mean absolute values reflecting overall impact. Individual Shapley values highlighted feature influence on correct classifications, which were also assessed for their relationships to psychopathology measures. FDR was used to control for false-positives in multiple comparisons. Correlational analysis between individual-level Shapley values for each feature and individual psychopathology. **Abbreviations:** Ada, AdaBoost; FDR, false discovery rate; RF, random forest; RSWG, Remission in Schizophrenia Working Group; SHAP, SHapley Additive exPlanations; SVM, support vector machine. EF, executive function.

## Results

### Participant identification, screening, and follow-up

In this hospital-based study, we initially identified 270 individuals with schizophrenia (International Classification of Diseases, tenth edition [ICD-10], additional screening with the Structured Clinical Interview for DSM-IV axis I Disorders) aged 18–65 years, from a total of 580 inpatients in the psychiatric department of the Third People’s Hospital of Lanzhou (Lanzhou, China) (Fig. 2). These 270 patients with schizophrenia were clinically stable (change in total PANSS score<20% with a particular type of antipsychotics at a maintenance dosage within the last 6 weeks; details in Supplementary Table S1). Furthermore, 75 of the 270 patients with schizophrenia were excluded due to illiteracy (*N*=35) or refusal to participate (*N*=40). Finally, 195 patients were included. The study was approved by the ethics committees of Northwest Normal University and the Third People’s Hospital of Lanzhou (Lanzhou, China). They underwent evaluations that included the electronic medical records, the Positive and Negative Syndrome Scale (PANSS), six EF behavioral tasks, and the fluid intelligence (Raven’s Progressive Matrices). To enable comparative analyses, 169 demographically matched healthy control participants who were free of a history of mental illness or brain injury were recruited and underwent the same assessments, except the PANSS.

**Fig. 2.**
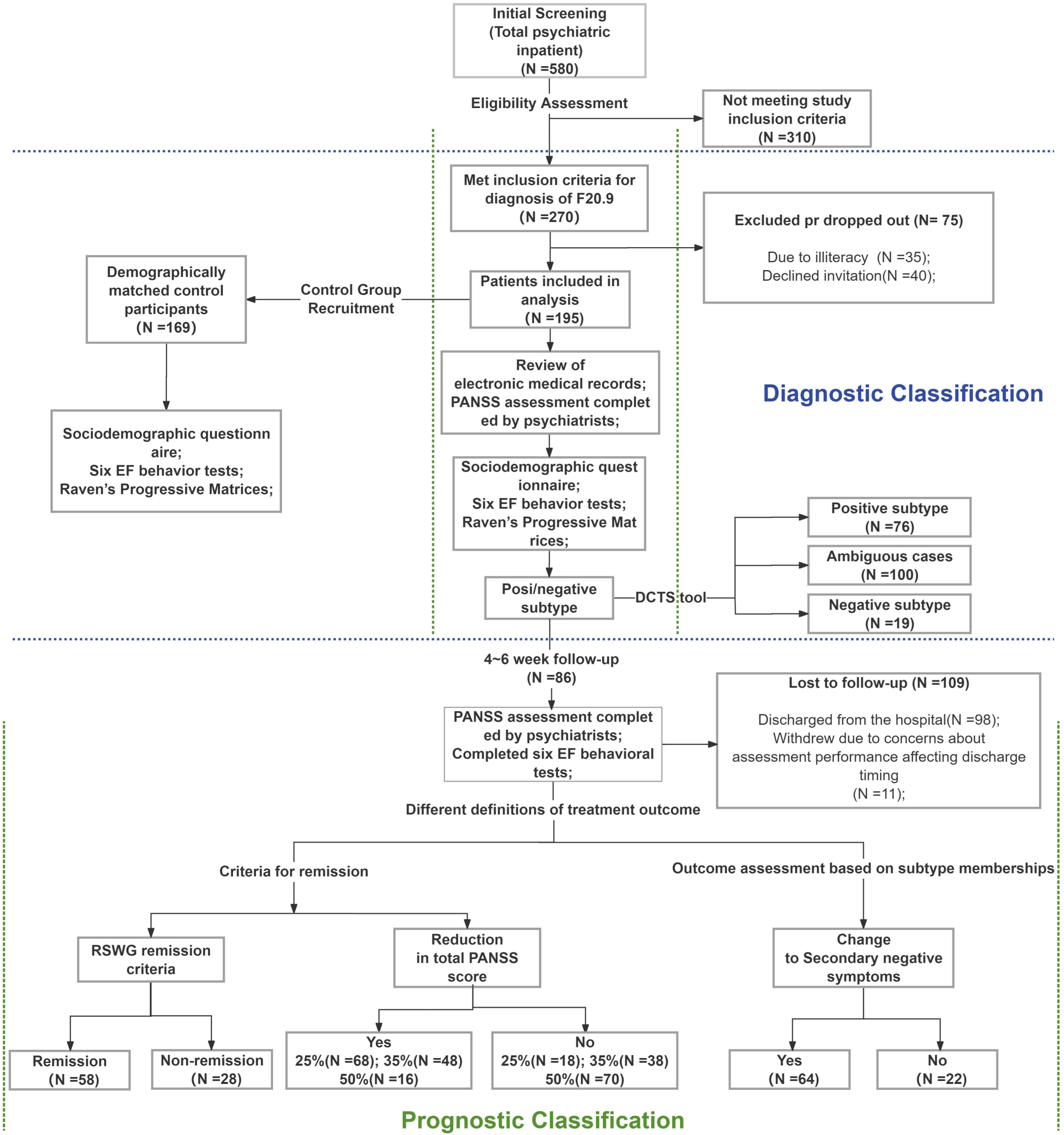
Flowchart of patient identification. Flow chart depicting the inclusion and exclusion criteria for participants. **Abbreviations:** DCTS, Dimensions and Clustering Tool for Schizophrenia Symptomatology; EF, executive function; PANSS, Positive and Negative Syndrome Scale; RSWG, Remission in Schizophrenia Working Group.

Among the 195 patients with schizophrenia, 86 participants completed follow-up assessments (PANSS and EF tests) in 4–6 weeks after the initial evaluation. The loss of 109 participants at follow-up was primarily attributed to hospital discharge (*N*=98) or concerns regarding the potential impact of assessment results on their discharge timing (*N*=11) (Supplementary Table S2). Fifty-eight participants at follow-up identified as being in remission; who had scores ≤ 3 on key PANSS items (P1, P2, P3, N1, N4, N6, G5, G9; the Remission in Schizophrenia Working Group [RSWG] criteria^34^). The other 28 participants were not in remission (Supplementary Table S2). Alternatively, defining the remission status by a reduction in PANSS total score at follow-up assessment relatively to baseline showed that^35–37^: 1) a 25% reduction (68 remitted vs. 18 non-remitted); 2) a 35% reduction (48 remitted vs. 38 non-remitted); 3) a 50% reduction (16 remitted vs. 70 non-remitted) (Supplementary Table S2). In addition, prognostic statuses may also be reflective in a change of subtype membership from baseline to the end of follow-up, particularly surrounds the negative symptom subtype. This is because many patients tend to experience increased negative symptoms and diminished positive symptoms due to standard antipsychotic treatments and related factors at follow-up, while others feature primary (stable) negative symptoms^38^ which relate to poorer clinical outcomes^39^. By using a subtyping system (http://webtools.inm7.de/sczDCTS/) from our previous work, patients with schizophrenia were assigned based on their symptom patterns as predominantly negative, positive, or ambiguous cases^20^. Of the 86 patients, 64 non-negative subtype patients (50 ambiguous and 14 positive) at baseline had a negative subtype assignment at follow-up assessment. Thirteen patients with schizophrenia maintained their negative subtype membership over time.

### EF dimensions

#### EF dimensions are consistently and differentially affected in schizophrenia

Six behavioral paradigms (e.g., Zhao et al., 2023)^40^, were administered to assess the three EF dimensions (inhibitory control, working memory maintenance and updating, and cognitive flexibility; Fig. 3A), based on 14 measurements: 1) The inhibitory control dimension: four measurements for the interference control function based on the *Stroop* task, three measurements for the response inhibition function based on the *Go/No-Go* task; 2) The working memory dimension: two measurements for working memory updating function based on the *running memory* task, three measurements for numeric working memory maintenance capacity based on the *Corsi block* test and the *digit span backward* task; 3) The cognitive flexibility dimension: two measurements (switch cost and mixing cost) in the *number-letter switching* task. Besides a conceptual formulation of the 14 task measurements according to the three-dimensional representation of EF, we supplemented five composite scores. This included an *Inhibition* composite score, an abbreviated version for representing general EF functions, and three cross-dimensional composite scores (*Inhibition/Switching, Inhibition/Working memory updating and Switching/Working memory updating*). Among these measurements, all reaction times,switching cost, and three composite scors (abbreviated general EFs, *Inhibition/Switching, Inhibition/Updating*) are the higher the worse EF performance, while the remaining the higher the better.

**Fig. 3.**
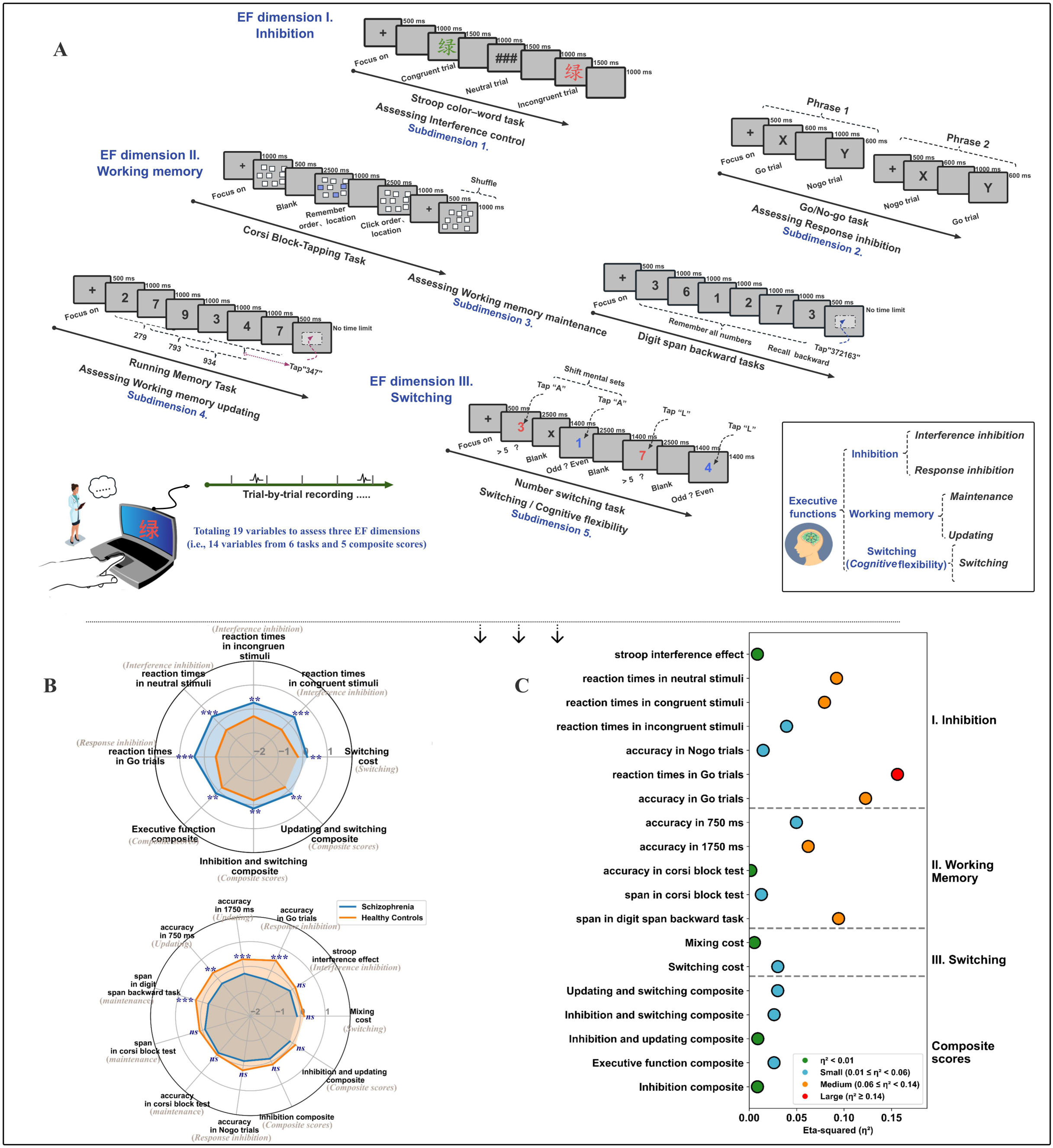
Multi-task executive function dimension assessments. **A).** The six behavioral paradigms (e.g., Zhao et al., 2023)^40^ used to assess the three EF dimensions (i.e., inhibitory control, working memory maintenance and updating, and cognitive flexibility) based on 14 measurements. **B)** The radar plot was constructed based on the Z-scores of the 14 EF measurements and the 5 composite scores, with annotated *p*-values (FDR corrected) resulted from one-way ANCOVAs following a mixed model ANCOVA. **C**). Effect sizes are colored as small (0.01 ≤ η² < 0.06), medium (0.06 ≤ η² < 0.14), or large (η² ≥ 0.14) based on the guidelines proposed by Cohen (1988). **Abbreviations:** EF, executive function; FDR, false discovery rate; *ns*, not significant. **Note:** *n*s: Not significant difference. *: *p* < 0.05. **: *p* < 0.01. ***: *p* < 0.001.

To evaluate whether schizophrenia differentially affected the three-dimensional EF measurements and the five EF composite scores, we performed a mixed-model analysis of covariance (ANCOVA). This has revealed a significant two-way interaction between group (schizophrenia vs. HC) and EF measurements (*p*<0.001). Follow-up one-way ANCOVAs revealed that, except for 2 measurements which assess working memory maintenance based on the *Corsi block* test, the accuracy in No-Go trails, the difference of reaction time between the congruent and the incongruent condition trials (i.e., the interference effect) in the Stroop task, the mixing cost in the switching task and the two composite scores of *Inhibition/Working memory updating* and *Inhibition*, other measurements and composite scores were all significantly different between patients with schziohrenia and healthy controls (all *p*<0.05, false discovery rate [FDR] corrected) (Table 2; Fig. 3B). These differed EF performance cover all of the three EF dimensions with the reaction times for Go trails (response inhibition) in the *Go/No-Go* task presenting the largest effect size (*η*^2^= 0.160) (Fig. 3C, Table 2).

#### Remitted patients show improved interference inhibition at follow-up, without difference in any EF dimension at baseline, compared with non-remitted patients

Further analyses were conducted to examine differences in baseline EF dimensions between the remission and non-remission groups (RSWG criteria); there were no significant between-group differences observed on any EF dimension (Supplementary Table S3). When comparing respective changes in these dimensions from baseline to follow-up (Supplementary Table S5), significant differences were only observed in the remission group. Specifically, on the Stroop task, the remission group demonstrated shorter reaction times under neutral, congruent, and incongruent conditions compared with baseline (i.e., better interference inhibition ability; all *p*<0.05).

Regarding clinical outcomes defined by subtype transmission, there were no significant differences in baseline EF measurements among the three subgroups (i.e., positive, negative, and ambiguous) of patients with schizophrenia (all *p*>0.05, FDR corrected). In patients with more prominent secondary negative symptoms (i.e., baseline non-negative subtype with transition to a negative subtype), the Stroop task revealed significantly reduced reaction times at follow-up compared with baseline for incongruent stimuli (*p*=0.022), congruent stimuli (*p*=0.007), and neutral stimuli (*p*=0.037). Additionally, the mixing cost in the switching task was significantly lower at follow-up (*p*=0.048). However, patients of the stable negative subtype during follow-up did not show significant differences between baseline and follow-up across all EF measurements. There were no significant differences observed in baseline EF measurements between the secondary negative group and the primary negative group (all *p*> 0.05).

### Psychopathology

#### Psychopathology dimensions specifically correlated with different EF measurements

Previous studies showed that EF, and specifically their dimensional measurements, could be associated with different aspects of symptomatology^41^. We probed the potential association of multifaceted aspects of psychopathology with different EF functions at the baseline assessment using Pearson correlation analysis (Supplementary Fig. S4). Using the four-dimensional structure of the PANSS^20^, we observed significant reductions in severity across negative, positive, cognitive, and affective symptoms in patients with schizophrenia at follow-up (*p*<0.001, Supplementary Table S6). Among the 86 follow-up patients, the ability to inhibit conflict, as reflected by reaction times to incongruent (*r*=0.212, *p*=0.050) and congruent stimuli (*r*=0.246, *p*=0.022) in the *Stroop* task, were significant correlated with baseline positive symptom scores. Patient capacity to maintain and shift mental sets, quantified by switch cost (*r*=0.298; *p*=0.005) and mixing cost (*r*=0.265; *p*=0.014) in the *number-letter switching* task, was significantly correlated with baseline cognitive and positive symptom scores, respectively, within the follow-up subset but not in the overall patient sample. For comparison, correlation analyses were repeated using PANSS three-original subscale scores; these yielded similar results, except for correlations with *Stroop* metrics (Supplementary Fig. S5).

#### EF dimensions show consistent and distinct impairments in schizophrenia and offer prognostic insights via machine learning classification models

Using a multivariate approach, classification models can be employed to identify feature variables (e.g., EF measurements) which can reliably differentiate target cases (i.e., patients with schizophrenia) from reference cases (i.e., HC). Additionally, they can provide insights into future categories (e.g., prognostic status) based on baseline assessments. Three methods (i.e., RF, SVM, and AdaBoost), and their stacked assembler, were used to construct classification models. The original data was repeatedly split into discovery and test sets, with each discovery set nested for hyperparameters tuning and model validation as a CV design. Next, the ensuing best model was applied to the test set from each repeat to obtain out-of-sample performance. The whole procedure was repeated for 100 tests, resulting in 100 hold-out, test sets. This approach has been demonstrated to effectively gauge generalization while balancing practical acquisitions of clinical sample data^42–44^.

##### 1) Diagnostic classification

For our classification experiments, two feature sets were used: 1) 19 EF assessments, which reflect three EF dimensions measured by six behavioral paradigms (Table 2); and 2) 32 features, which added 13 sociodemographic variables to the 19 EF variables in feature set 1 (Table 2). Performance metrics were assessed on the 100 test sets (Supplementary Table S7). For the feature set relying on only EF assessments (i.e., feature set 1), we aimed for a model to classify new patients, regardless of sociodemographics. The highest out-of-sample classification was achieved by the stacking model (area under the curve [AUC]=0.87). The feature set 2, which also included sociodemographic variables, was aimed at incorporating information from routine clinical interviews that indicates disease susceptibility^45^. With the addition of sociodemographic variables, improved model performance was observed (stacking model AUC=0.91). In addition, repeating the entire CV process on the EF assessments after controlling for the effects of sociodemographic variables largely maintained the performance (stacking model AUC=0.80) (Fig. 4A).

**Fig. 4.**
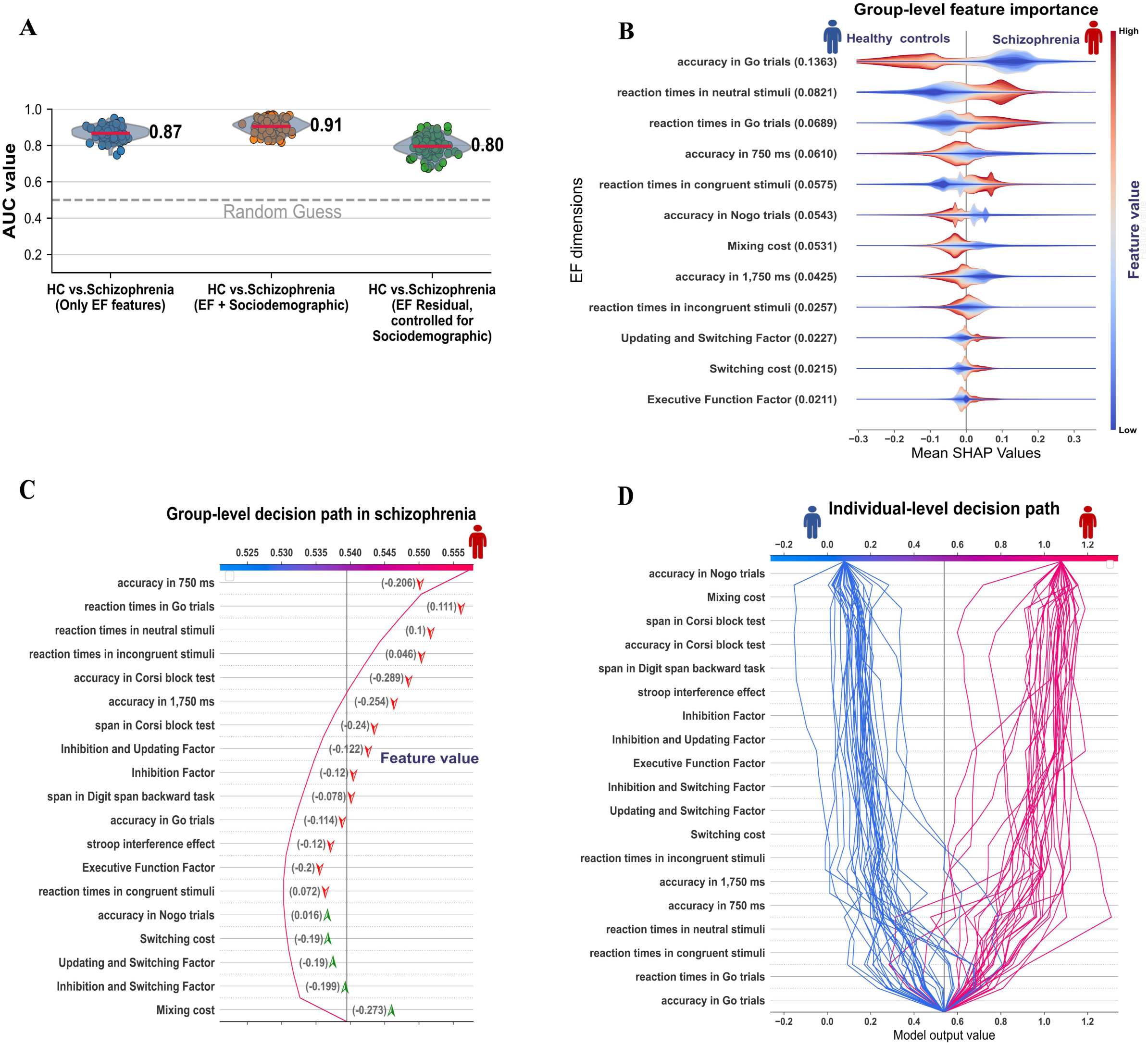
Model metrics and feature importance for diagnostic models. **A).** Violin plots show the values of area under the curve (AUC) for discriminating patients with schizophrenia from healthy participants by diagnostic classification models. Each point within the violin plots represents the AUC value derived from the hold-out test data of each random split procedure (repeated 100 times) in our machine learning design. The red line within each violin plot denotes the mean. **B).** The group-level feature importance plot ranks EF features on the y-axis by their absolute average Shapley value across individuals, representing their overall importance in the ability of the model to distinguish between patients and healthy participants. The original feature weights for each EF variable were color coded, with blue color denoting a negative weight value and red color denoting a positive weight value. Values along the x-axis indicate a positive or negative effect of an EF feature on classifying an individual, with a negative and positive value promoting the model towards a classification of "healthy" and “schizophrenia”, respectively. Collectively, these findings indicated that higher accuracy in the Go trials was associated with a higher likelihood of classifying an individual as healthy. **C).** The group-level decision path, generated based on all correctly classified schizophrenia samples. EF features are ranked from upper to bottom based on their group-level importance present in b), the color bar denotes the impact of an EF feature on model’s classification towards “healthy” (blue) or “schizophrenia” (red), the red curve shows the values for each EF feature coded in the color bar. The numbers in parentheses represent the z-score standardized original measurement values of each EF feature by the averages of this EF feature across the healthy and the patient groups in the model test samples (a negative number to the right of the perpendicular line denotes the measurement of an EF feature that is below the average in patients with schizophrenia). The red or green arrows adjacent to the parentheses indicate the higher or lower values within the parentheses that are associated with better or worse EF functions, respectively. This is because some measurements from the EF tasks indicate better performance with higher values, while others are more favorable with lower values. **D).** The decision path for each individual in the test sets of the classification modeling iterations, that for each individual, how each of these important EF features have promoted the diagnostic model to classify this individual as a healthy participant or patient with schizophrenia, given the expression level (task measurements) of this individual in each of the EF features. The blue and red lines indicate accurate classifications as healthy participants and schizophrenia patients, respectively. **Abbreviations:** EF, executive function; HC, healthy control.

##### 2) Prognostic classification

For prognostic classification, we applied the same models (RF, SVM, AdaBoost, and their stacking, as described in the diagnostic classification above) on two feature sets, to assess their performance in discriminating remission status: (1) 19 baseline EF variables; (2) 32 features (the 19 baseline EF variables plus 13 sociodemographic variables). The stacking model achieved the highest classification accuracy based on the EF assessments-only feature set (AUC=0.81), and the performance was identical to that observed when the EF plus sociodemographic features set was used.

Additionally, to test the influence of demographic variables and medication on our models, we conducted several control analyses (Fig. 5A). By regressing out the effects of sociodemographic variables on EF assessments, we found that the model performance was decreased to AUC=0.65, indicating a poorer setup (Fig. 5A). However, there was no significant difference in any of the sociodemographic variables adjusted in our classification models between the remitted and non-remitted patient subgroups (all *p*>0.05) (Supplementary Table S3). Additionally, controlling for medication effects using an olanzapine (OZP) equivalent dosage (which did not differ significantly between remitted and non-remitted patients; *p*=0.15) diminished the prognostic classification accuracy to an AUC of 0.72. By investigating only those patients (*N*=46) treated with a commonly effective OZP-equivalent dosage of 10–20 mg/day in clinical practice produced a similar prognostic classification performance (AUC=0.82) to that recorded in the group of followed up patients (*N*=86).

**Fig. 5.**
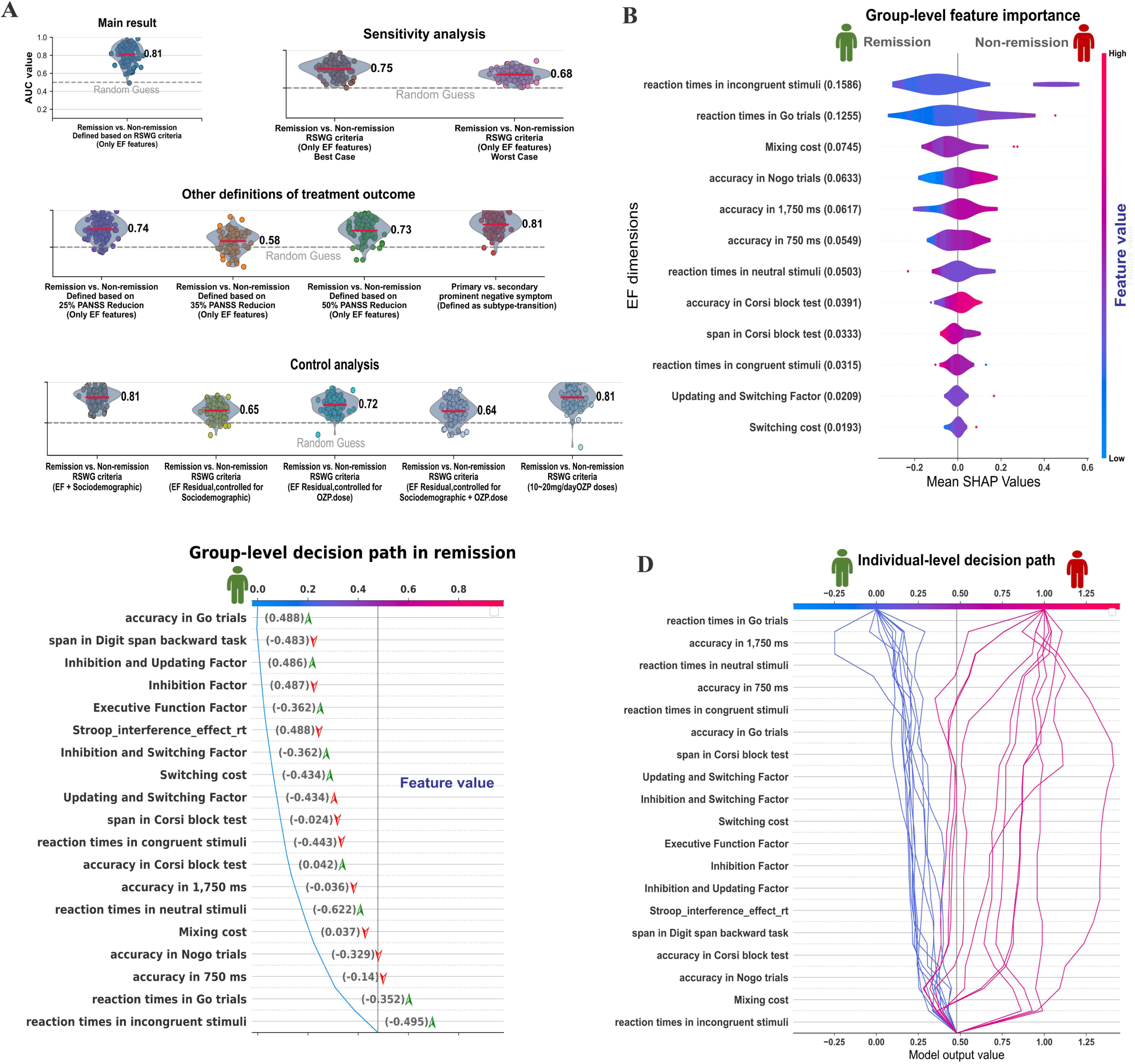
Model metrics and feature importance for prognostic models. **A).** Violin plots show the values of area under the curve (AUC) for discriminating remitted patients with schizophrenia from non-remitted patients by prognostic classification models. Each point within the violin plots represents the AUC value derived from the hold-out test data of each random split procedure (repeated 100 times) in our machine learning design. The red line within each violin plot denotes the mean. **B).** The group-level feature importance plot ranks EF features on the y-axis by their absolute average Shapley value across individuals, representing their overall importance in the model’s distinction between remitted and non-remitted patients (i.e., greater separation of the violin-like plots towards the extremes denotes higher importance). The original feature weights for each EF variable are color-coded, with blue indicating a negative weight value and red indicating a positive weight value. Values along the x-axis indicate a positive or negative effect of an EF feature on classifying an individual, with a negative and positive value promoting the model towards a classification of "remission" and “non-remission”, respectively. Collectively, these data suggest that longer reaction times in incongruent stimuli are associated with a higher likelihood of classification as non-remitted patient. **C).** The group-level decision path, generated based on all correctly classified remitted schizophrenia patients. EF features are ranked from upper to bottom based on their group-level importance present in b), the color bar denotes the impact of an EF feature on model’s classification towards “remission” (blue) or “non-remission” (red), and the blue curve shows the values for each EF feature coded in the color bar. The numbers in parentheses represent the z-score standardized original measurement values of each EF feature by the averages of this EF feature across the remitted and non-remitted patient subgroups in the model test samples (a negative number to the left of the perpendicular line denotes the measurement of an EF feature that is below the average in remitted patients). The red or green arrows adjacent to the parentheses indicate the higher or lower values within the parentheses that are associated with better or worse EF functions, respectively, as some measurements from the EF tasks indicate better performance with higher values, while others are more favorable with lower values. **D).** The decision path for each individual in the test sets of the classification modeling iterations, illustrating how each of these important EF features influenced the prognostic model to classify each individual as remitted or non-remitted, given their performance (task measurements) in each of the EF features. The blue and red lines represent correct classifications as remitted and non-remitted patients, respectively. **Abbreviations:** EF, executive function; OZP, olanzapine; PANSS, Positive and Negative Syndrome Scale; RSWG, Remission in Schizophrenia Working Group; SHAP, SHapley Additive exPlanations.

We supplemented classifications to discriminate prognostic subgroups defined by the reduction in re-assessed total PANSS score. The results showed a poorer discriminative power, with AUCs of 0.74, 0.58, and 0.73 (Fig. 5A; Supplementary Table S8) for a 25%, 35%, and 50% reduction in the PANSS score, respectively. Alternatively, we established a classification model for the clinical outcomes of patients based on subtype-membership transition to distinguish between 1) baseline non-negative subtype with a transition to negative subtype (secondary) and 2) stable negative subtype during follow-up (primary). A promising classification performance was revealed (AUC=0.81).

Moreover, two sensitive analyses were performed to take the attribution condition in our study into consideration. The demographic variables, symptoms, OZP-equivalent dosage, and EF measurements did not differ significantly between patients who were followed up and those who were not (all *p*>0.05, Supplementary Table S3). Furthermore, by treating the attribution patients as best cases (i.e., all remitted), the classification accuracy decreased slightly to an AUC of 0.75, while treating the attribution patients as worst cases (none remitted) yielded a further decease (AUC=0.68).

#### Feature importance and association with individual psychopathology

SHAP analysis was performed on the best-performing classifiers trained on the 19 EF dimension assessments (for the feature sets including sociodemographic variables please refer to Supplementary Table S9)^46^. The goal was to determine the directional contribution of EF dimensions for classification informed by decision path (i.e., better or worse performance in an EF dimension drives the model to assign a schizophrenia or HC label). Besides group-level determination of important dimensions, the Shapley value for each EF feature in the best-performing classifiers was calculated for each individual^47,48^, facilitating a link to the expression level of patients along several psychopathological dimensions.

##### 1) Group-level feature importance and decision path

Absolute Shapley values derived from the best-performing classifier (highest AUC) identified in the 100 test sets were used to rank each EF dimension to indicate its importance in discriminating patients with schizophrenia, and those remitted at follow-up (Figs. 4B, 5B). Including only the 19 EF dimensions scores as the feature set in both diagnostic and prognostic classifications, the inhibition control dimension—comprised of response and interference inhibitions—ranked highest in both classifying schizophrenia group participants and their follow-up remission status. Within the inhibition control dimension, important features were from the *Go/No-Go task*, which assesses response inhibition (Go trial accuracy and reaction time), and the Stroop task, which assesses interference inhibition (reaction time for neutral stimuli in the diagnostic model, and reaction time for incongruent stimuli in the prognostic model). Adding sociodemographic variables to the diagnostic and prognostic classifiers generally replicated inhibition control as the strongest contributing dimension (Supplementary Figs S6A, S7A).

Next, group-level decision path analysis of each EF dimension identified that poor performance on any of these drove the model to correctly classify individuals as patients with schizophrenia (Fig. 4C). However, worse performance on the inhibition control dimension increased the likelihood of non-remission status classification (Fig. 5C). These results were replicated in additional models in which sociodemographic variables were included in diagnostic and prognostic classifiers (Supplementary Figs. S6B, S7B).

##### 2) Individual-level decision path and association with psychopathology

Decision path analysis was likewise conducted at the individual level, to determine the relative performance of EF dimensions (as identified in group averages) for correctly assigning individuals. Among correctly identified participants, patients with schizophrenia scored below the averages of both HC and schizophrenia groups on at least one EF dimension (Fig. 4D). As expected for the prognostic classification model, remitted status for most participants in the schizophrenia group was correctly assigned based on higher baseline inhibitory control dimension (including both interference control and response inhibition) performance compared with the averages of both remitted and non-remitted participants. Specifically, remitted participants showed shorter reaction times to the incongruent condition in the *Stroop* task, and higher accuracy in the response to the Go trials during the *Go/No-Go* task. However, a few patients presented both worse performance in inhibitory control and higher baseline abilities in other dimensions such as working memory updating or shifting (Fig. 5D; Supplementary Fig. S9). These findings were replicated when sociodemographic variables were included in diagnostic classification models (Supplementary Materials).

Pearson correlation analysis was further performed on Shapley values for each EF feature and scores on the four symptom dimensions, across the overall schizophrenia group (*N*=195) and follow-up subset (*N*=86). Results showed that individual Shapley values of the interference inhibition function, as assessed by a difference in reaction time between the congruent and the incongruent trials in the *Stroop* task, that promoted a correct assignment of cases versus HC were significantly associated with the negative symptoms (*r*=0.439, *p*=0.042, FDR corrected) for individuals with schizophrenia. The inference inhibition function, though assessed by a different behavioral metric, was similarly identified as a factor of top importance at the group-level for the diagnostic model described above (Fig. 6A). After including the 13 sociodemographic variables, there was no significant correlation found.

**Fig. 6.**
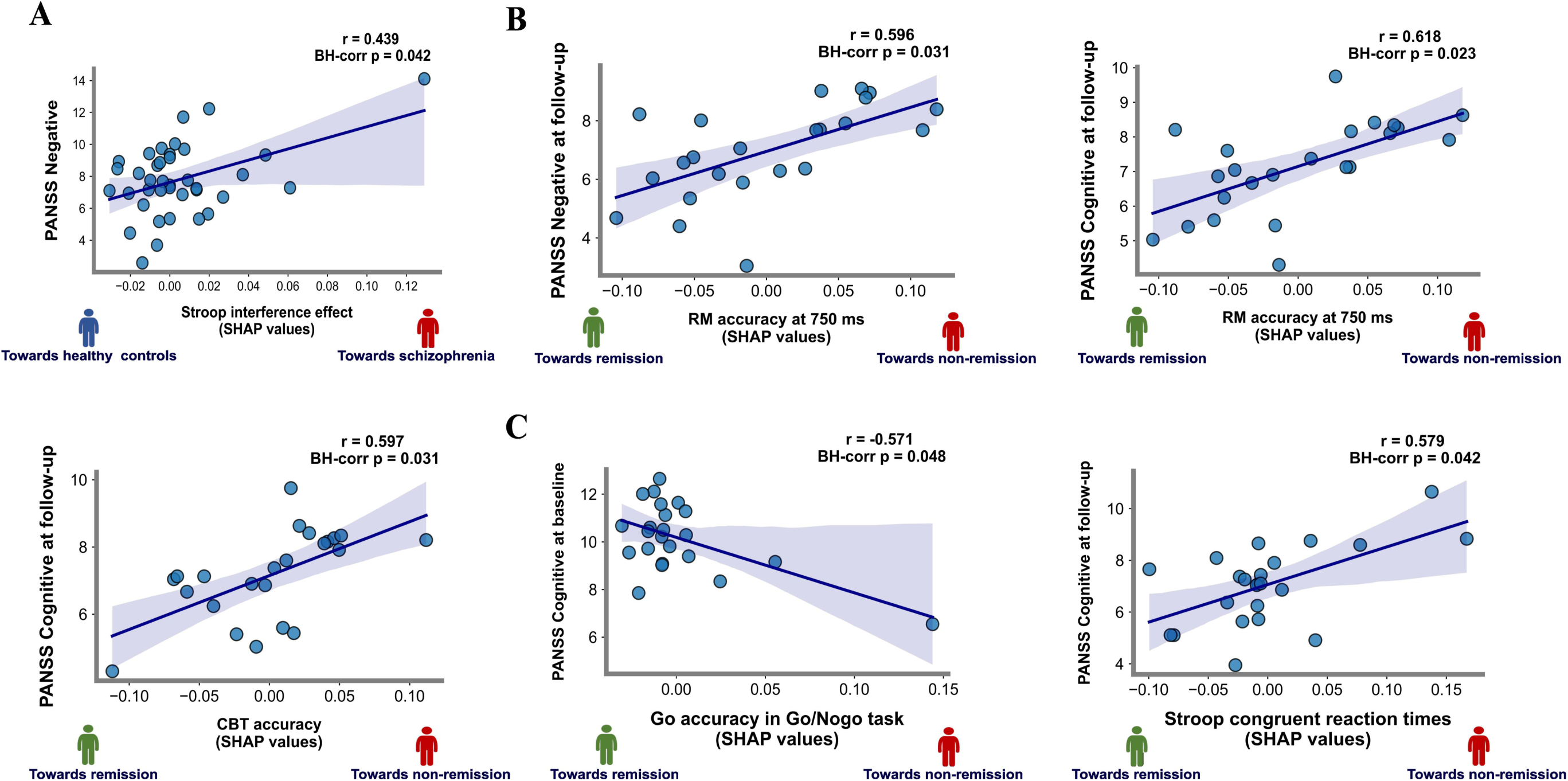
Correlation between the importance of an executive function feature and individual psychopathology along four symptom dimensions. **A).** Correlation in the diagnostic model using executive function features only. **B).** Correlation in the prognostic model using executive function features only. **C).** Correlation in the prognostic model using executive function features and sociodemographic features. **Abbreviations:** BH, Benjamini–Hochberg correction; CBT, Corsi block test, assessing numeric working memory maintenance capacity; Go/No-Go task, assessing response inhibition function; PANSS, Positive and Negative Syndrome Scale; RM, running memory task, assessing working memory updating capability; SHAP, SHapley Additive exPlanations; Stroop, Stroop task, assessing interference control function.

For prognostic classification, the importance of working memory updating (assessed in the *running memory* task; *r*=0.618, *p*=0.023, FDR corrected) and maintenance (assessed in the *Corsi block* test; *r*=0.597, *p*= 0.031, FDR corrected) in the model that accurately assigned remitted patients was correlated with low-level cognitive symptoms at follow-up. Additionally, working memory updating function contributing to the accurate assignment of remitted patients was associated with more severe re-assessed negative symptoms (*r*=0.596, *p*=0.031, FDR correction). Following the inclusion of sociodemographic variables in the model, the significantly associated EF variables were changed (Fig. 6C).

Using PANSS three-subscale scores in correlation analyses did not reveal significant correlation with the importance scores of any EF features of the dimensions identified in the diagnostic models. Significant correlation patterns among the importance scores of EF dimension features in prognostic models were a subset of those reported (Supplementary Fig. S10) when using the four dimensions of psychopathology, as described above.

## Discussion

This study is the first to investigate the classification power of comprehensively three EF dimensions (i.e., inhibitory control, working memory maintenance and updating, cognitive flexibility) for discriminating both patients with schizophrenia from HC and determine their remission status at follow-up. Importantly, our SHAP approach parsed the relative importance of each feature in these classification tasks, at the group and individual level. Collectively, we found that EF dimensions consistently and differentially characterize schizophrenia and are informative regarding the prognostic status, though certain dimensions are more closely linked to the disease trait and related psychopathology.

The four primary findings are as follows. Firstly, EF assessments could be used to both classify patients with schizophrenia (AUC=0.87) and to identify those with remitted status (AUC=0.81). Importantly, there is no significant difference in baseline EF and symptom severity between the two prognostic subgroups. Secondly, inhibition control was the most strongly contributing dimension to patient classification of both schizophrenia and remission outcome. Thirdly, at the individual level, correctly identified patients presented below-average performance on at least one EF dimension. However, except for a few patients with correctly assigned remission status who had worse performance in inhibitory control, others featured higher baseline performance in this dimension compared with the averages of both remitted and non-remitted patients. Finally, the EF dimension interference inhibition, which is important in promoting the correct classification of patients with schizophrenia by the model, was significantly associated with patient negative symptoms. The model importance of working memory for accurate remission assignment covaried with low-level follow-up cognitive symptoms.

The paradigms measuring EF and its dimensions are readily available through software platforms, such as *E*-Prime and Matlab-based Psychtoolbox. Thus, they can be implemented in clinical practice. The present findings may encourage the use of a feasible, low-cost, and effective approach to schizophrenia diagnosis and psychopathology evaluation.

### Schizophrenia, and its remission status, are classified by EF dimensions

Previous machine learning studies employing neuropsychological test batteries (e.g., the Cambridge neuropsychological test automated battery, the Wechsler adult intelligence scale, and the brief assessment of cognition in schizophrenia) to differentiate patients with schizophrenia from HC have yielded accuracy rates <70%^8,49^. Neuroimaging-derived assessments are an alternative and broadly attempted approach^50^. While showing promise for improving classification performance^51,52^, this strategy is associated with other challenges^53^ including heterogeneous data acquisition, high dimensionality due to large numbers of voxels or measures, and limited applicability in low-income countries and regions, as illustrated by our recent meta-analysis of global psychiatric neuroimaging data^54^. Another direction is systematic modeling based on readily attenable data with improved objectivity and reliability, namely behavioral EF tasks, for improved clinical translation^10,55^. By carefully assessing three EF dimensions via six tasks, and thus 19 variables, our stacking model achieved reasonably high diagnostic accuracy. Furthermore, the inclusion of sociodemographic characteristics increased the AUC to 0.91. This is broadly consistent with previous findings of an association between sociodemographics and disease susceptibility, clinical course, and symptom expression in schizophrenia^45^. In clinical practice, early and accurate prediction of remission outcomes holds significant implications for effective treatments. Nevertheless, this remains difficult, with recent models based on baseline neuroimaging, genetics, and clinical factors often producing accuracy rates marginally better than the chance level (i.e., 50%). Comparatively, our prognosis classification model, incorporating only baseline EF assessments, demonstrated improved performance in denoting remission status among patients with schizophrenia at 4–6-week follow-up (AUC=0.81). We moreover tested the classification models by two methods for defining remission, i.e., based on a reduction in re-assessed PANSS scores calculated with specific items or all items, to assess the robustness of our findings to the definition of treatment response^36^. The discriminative power of the classification model was higher when specific items, compared with all items, of the PANSS were involved in defining prognostic statuses. This evidence implied specificities in mapping EF dimensions and symptom recovery in patients with schizophrenia.

### Classification-important EF dimensions and associations with individual psychopathology

Leveraging the SHAP framework, we conducted feature importance analysis on our classification models to identify the relative contributions of each EF dimension variable.

### Diagnostic classification

At the group level, we found that the inhibitory control dimension (i.e., interference inhibition and response inhibition) ranked highest in importance for classifying participants in the schizophrenia group. This is consistent with previous works showing abnormal alterations in the temporal and spatial characteristics of inhibition-related brain responses and behaviors in schizophrenia^56,57^. Moreover, our decision path plot pertaining to all correctly classified individuals indicated that those who performed poorly on any EF dimension tended to be classified in the schizophrenia group. This emphasizes the general EF impairments among patients with schizophrenia, consistent with a previous meta-analysis showing that EF deficits within this patient population cover broad dimensions^26^. Considering individual variation in this context through decision path analysis for each participant, we noted that a few accurately identified patients with schizophrenia performed slightly above average on either of the three EF dimensions (e.g., inhibition, updating, or shifting). This aligns well with data showing that cognitive performance, including EF functions, in schizophrenia can vary from mild deficiency^58,59^ to severely impaired^60,61^, and connects the neuropsychological and neurobiological heterogeneity systematically observed among these patients^62,63^.

By extending the SHAP framework to individual-level analyses, we further linked the importance of each EF feature in diagnostic classifier—quantified by Shapley values—to the landscape of individual psychopathology. Notably, we found Shapley values from the dimension identified as important at the group-level—interference inhibition—to be valuable within the model correctly classifying patients with schizophrenia, and significantly associated with a patient’s negative symptom expression. Consistent with our finding, earlier research showed that individuals with more severe negative symptoms tend to have diminished inhibitory control, as measured by the *Stroop* task^64^. This deficiency, particularly within interference inhibition, might play a role in the manifestation of negative symptoms, which may be understood through the lens of target-speech recognition deficit in schizophrenia^64^. Specifically, the interference inhibitory function assists individuals in accurately segregating target speech from noisy background environments in which multiple speakers are talking simultaneously^65^. An impairment in this function would thus be connected to the disorganized speech information processes in schizophrenia, including the inability to either inhibit unrelated speech signals or capture desired speech signals^65^. Such impairment has been correlated with the severity of negative symptoms, including poverty of speech and hypobulia^66^.

### Prognostic classification

Interestingly, while inhibitory control contributed to diagnostic classification, it was also the top predictor of remission outcome at follow-up. Our decision path analysis showed that patients who were correctly classified as remitted generally (despite some exceptions) showed good baseline performance on inhibitory control tasks. Previous work has consistently demonstrated an association between EF and long-term post-treatment remission outcomes in patients with schizophrenia^67^. Specifically, patients with higher EF performance are more likely to remit relative to those with lower EF performance^68,69^, especially on the inhibition control dimension^70^. A possible interpretation is that patients with better inhibitory control are more adherent to therapeutic plans, including pharmacological interventions and lifestyle modifications^71,72^. Alternatively, because inhibitory control assessments are closely related to specific clinical manifestations in patients with schizophrenia^73–75^, better performance on this EF dimension along with milder symptom expressions among such patients, implies higher chance of remission^68^. Our results showed significant post-treatment improvement in interference inhibition in the remission subgroup, but not in the non-remission subgroup (Supplementary Materials), corroborating a relationship between inhibitory performance and remission in schizophrenia. Interestingly, we did not find significant difference in any baseline EF assessment and symptom dimension score between remitted and non-remitted patients. This points to a dissociation between symptoms of schizophrenia and the construct of EF; nonetheless, it highlights the role of EF dimensions in predicting remission rather than merely acting as a marker of illness severity.

A few patients who were correctly classified as having symptom remission exhibited poor inhibitory control performance; nevertheless, they demonstrated better baseline abilities in other dimensions, such as working memory updating or shifting. Furthermore, we revealed that the importance of working memory updating and maintenance contributions to the prognostic model in classifying remission status covaried with poor patient cognition at follow-up. Supporting this observation, previous studies have shown that working memory (updating and maintenance) performance is superior among stably remitted patients with schizophrenia versus non-remitted patients^68^. Disrupted working memory in patients with schizophrenia has been linked to cognitive disorganization and poorer performance on tasks requiring abstract thinking^24,76,77^, which is similarly assessed within our cognitive symptom dimension based on PANSS.

### Limitations and considerations

First, applied machine learning tends to rely on multiple datasets from independent medical centers for extensive tests. However, such concerns are moderated by our use of multiple random-splits to set aside a test ‘lock box’ in each repeat, while performing nested CVs on the remaining sample, as well-established previously^50,53^. This strategy effectively gauges the out-of-sample generalization performance, while balancing practical clinical data collection issues^42,43,78,79^. Nevertheless, future multisite and population-level EF studies may help expand the applicability. Second, patients with schizophrenia in our study had been treated with antipsychotics, reflecting typical clinical practice. In our sample, the OZP equivalent dosage did not show a significant correlation with most symptom scores (except for the affective symptom dimension) or EF measurements (except for one variable). Moreover, there was no significant difference in baseline OZP dosage between patients who were in remission and those who were not at follow-up. Nevertheless, the exact dosage of antipsychotic medication could have an impact on individual prognostic status^80^. As expected, the classification accuracy decreased when adjusting for individual variations in OZP equivalent dosage. However, a control analysis that only included patients who received the suggested starting dose for OZP (e.g., 10–20 mg/day)—a dose commonly effective in individuals with schizophrenia^81^—remained the classification performance as in our main experiments. Notwithstanding, future research involving drug-naïve patients at baseline and continuous assessments of EF function along with detailed records of medication usage over time would help establish the causal relationships between antipsychotic effects, prognostic statuses, and specific EF dimensions. Third, the attrition rate in our patient sample was similar to those reported previously.^82–84^ This rate pertains to the representativeness of the findings derived from the patients who continued in the study, though we did not observe significant differences in all of the baseline characteristics between the followed up and drop-out patients (Supplementary Table S3). Furthermore, classification models established based on the two extreme conditions (i.e., treating the attribution patients as best-case [all remitted] or worst-case [non-remitted] scenarios) showed decreased prognostic discrimination accuracy. This was in line with the previous notion^85,86^ that the worst case analysis would lead to underestimated results^87^, though these might not reflect the true potential attrition bias. Future research may also incorporate outpatients to develop models representing the broader spectrum of patient populations, but managing the attrition rates remains a challenge.

To conclude, here we tested the classification power of six well-established behavioral paradigms, which assess three EF dimensions, for discriminating patients with schizophrenia from HC at baseline, as well as the remission status at follow-up. Results from robust validation and testing revealed promising performance and, thus, a consistent impairment in dimensions of EF to characterize individuals with schizophrenia and provide important prognostic information. Furthermore, different EF dimensions characterized diagnosis and prognosis to varying extents. Inhibitory control and working memory were identified as the most important factors for accurate classification of schizophrenia and remission status. Additionally, the classification strength of these EF dimension features was associated with specific psychopathologies. Thus, our research presents evidence that certain dimensions of EF are reliably compromised in individuals with schizophrenia. These deficits correlate with the severity of symptoms and can predict future outcomes. Hence, these measures may serve as valuable aids in the clinical assessment of this disorder.

## Methods

### Ethics approval and consent

This study was conducted in full compliance with the ethical guidelines and approved protocols of the Ethics Committee at the Third People’s Hospital of Lanzhou City and Northwest Normal University (Lanzhou, China). Before participation, all participants involved in the study were provided with comprehensive information regarding the study aims, procedures, potential risks, and benefits. The study adhered to the principles outlined in the Declaration of Helsinki, and informed consent was provided by all participants.

### Data collection

#### Participant recruitment, clinical characterization, and definition of prognostic status

The study was conducted from March to August, 2023. Participants were 195 individuals who had been diagnosed with schizophrenia (of those, 86 were further assessed after a 4–6-week interval), and 169 healthy individuals (HC group) (Table 1). Participants in the schizophrenia group had received inpatient treatment at the Third People’s Hospital in Lanzhou City within the past 2 years. Diagnoses were reached by two resident psychiatrists using the ICD-10 diagnostic criteria for schizophrenia (F20.9). The participants were further screened with the Structured Clinical Interview for DSM-IV axis I Disorders. The patients were in a stable condition and receiving consistent treatment, with no changes in medication expected during the study. Inclusion criteria were age 18–65 years and the ability to communicate effectively, complete experimental tasks, and voluntarily sign the informed consent form. Individuals with a severe physical disease, visual abnormality, or adverse drug reactions were excluded from the study (details for the inclusion and exclusion criteria are listed in Supplementary Table S1). Participants in the HC group, recruited through offline promotions and online advertisements, were matched with those in the schizophrenia group for age, sex, education level, and socioeconomic status. All HC group participants were physically healthy and did not have a history of mental illness or brain injury.

**Table 1.**
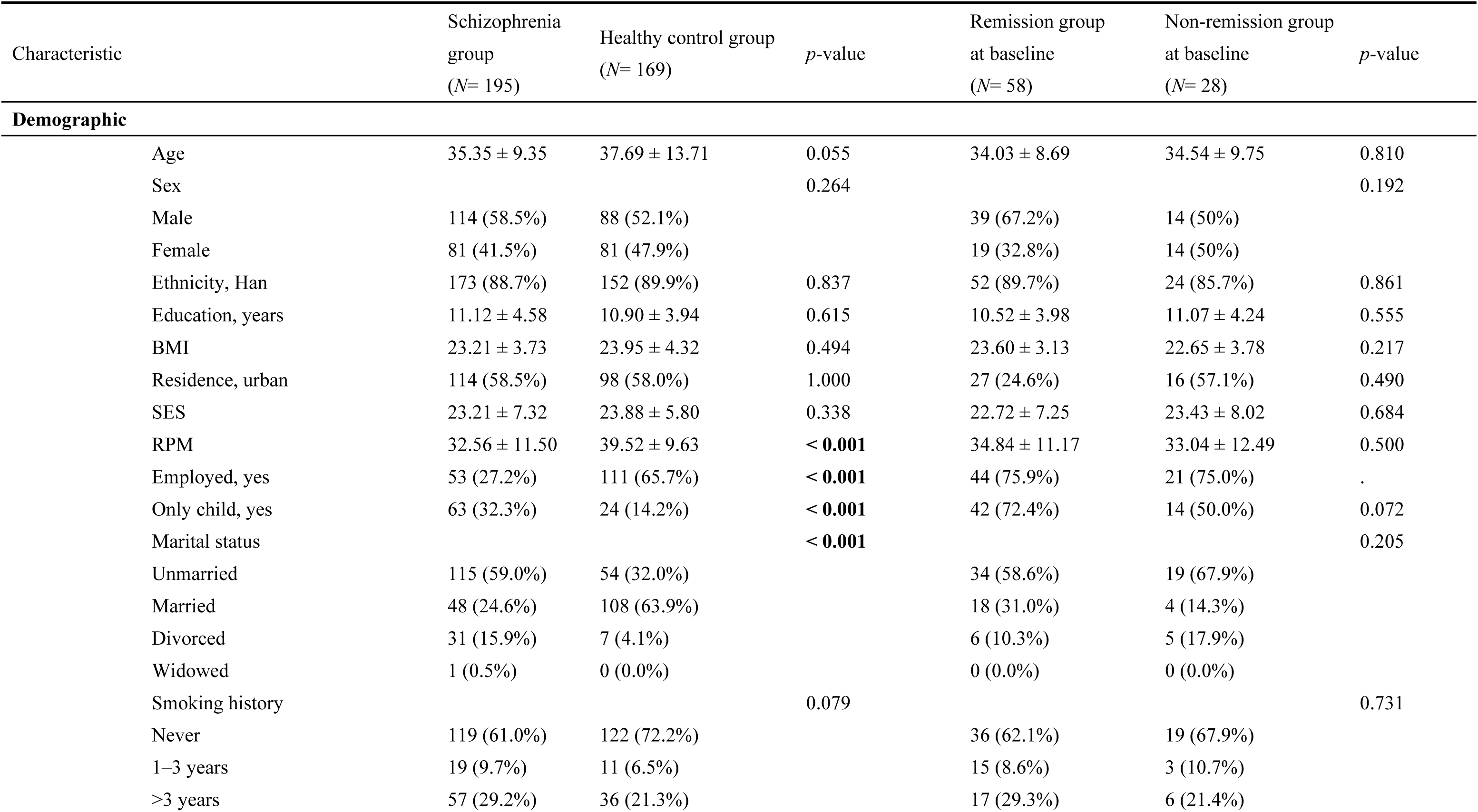
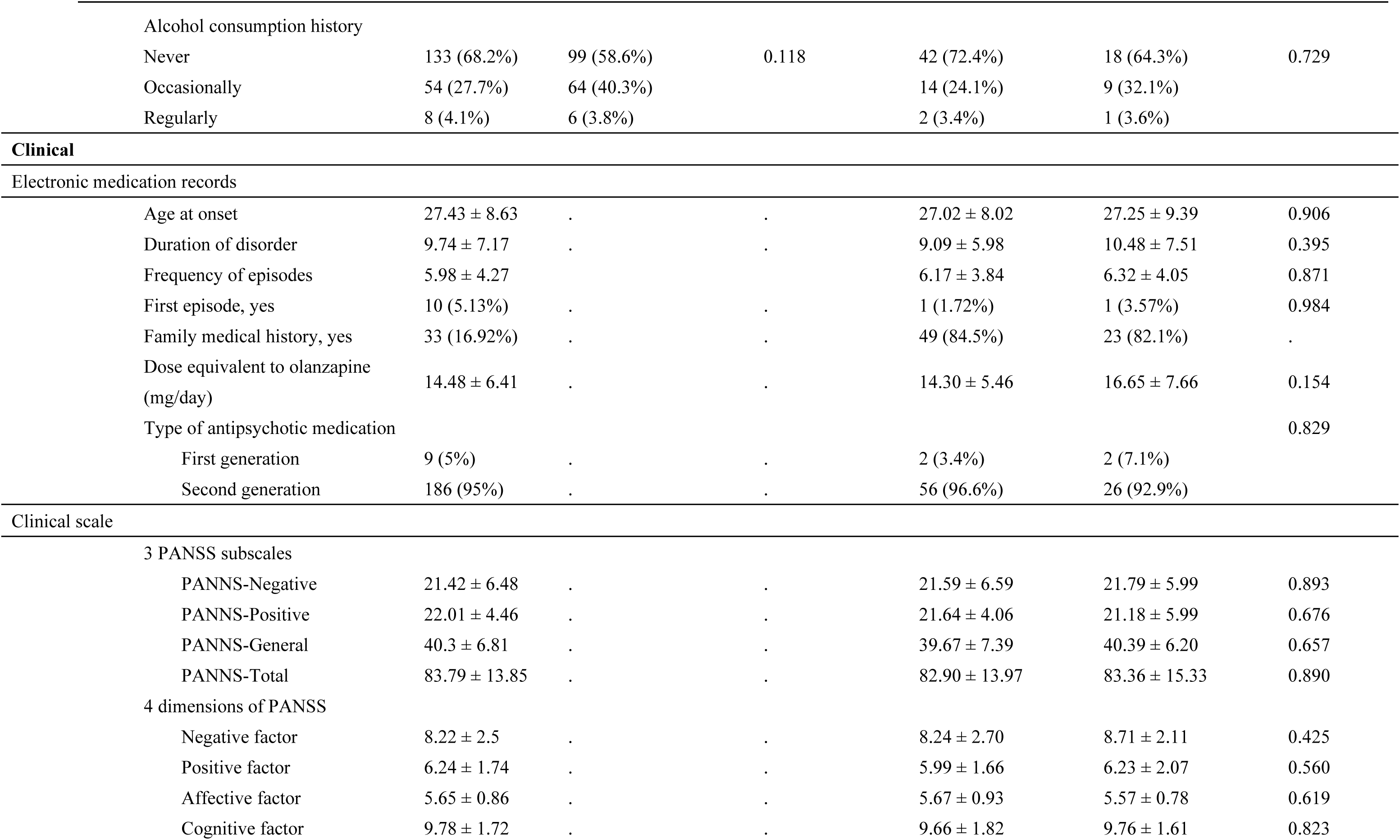
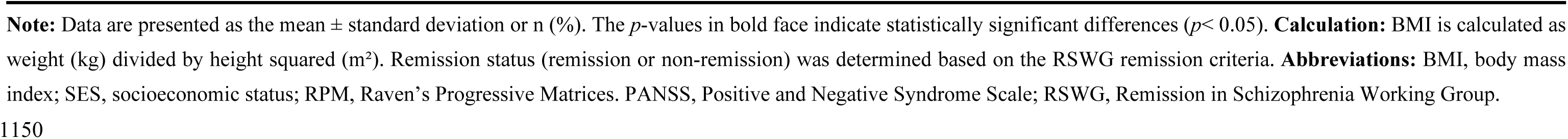
Participant demographics and clinical characteristics.

**Table 2.**
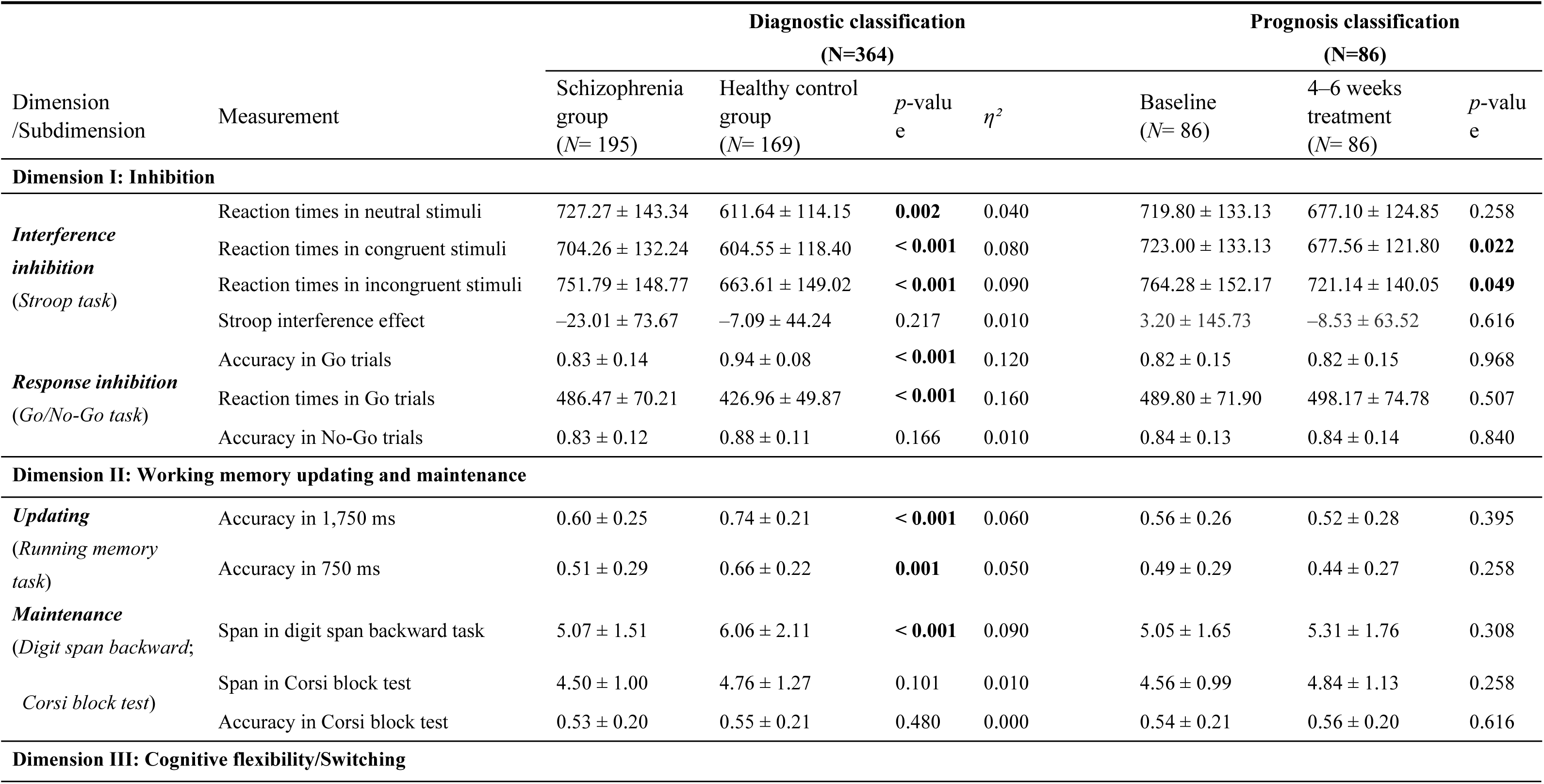
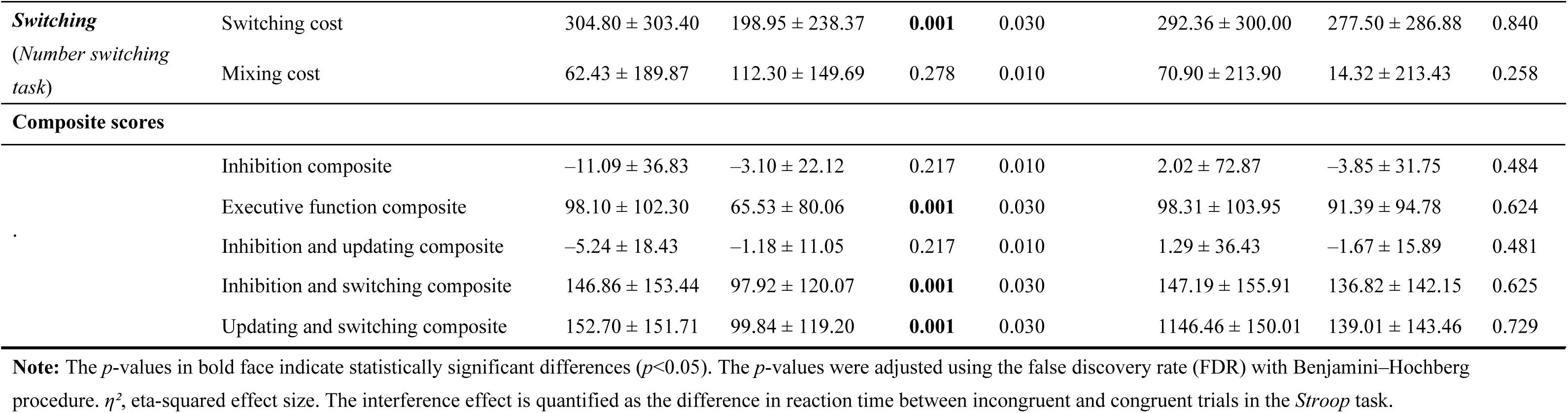
Comparisons of executive function dimensions between groups.

|  |  | Diagnostic classification |  |  |  | Prognosis classification |  |  |
| --- | --- | --- | --- | --- | --- | --- | --- | --- |
|  |  | (N=364) |  |  |  | (N=86) |  |  |
| Dimension /Subdimension | Measurement | Schizophrenia group<br>*(N= 195)* | Healthy control group<br>*(N= 169)* | *p*-valu e | $\eta^2$ | Baseline<br>*(N= 86)* | 4–6 weeks treatment<br>*(N= 86)* | *p*-valu e |
| **Dimension I: Inhibition** |  |  |  |  |  |  |  |  |
| ***Interference inhibition***<br>*(Stroop task)* | Reaction times in neutral stimuli | 727.27 ± 143.34 | 611.64 ± 114.15 | **0.002** | 0.040 | 719.80 ± 133.13 | 677.10 ± 124.85 | 0.258 |
|  | Reaction times in congruent stimuli | 704.26 ± 132.24 | 604.55 ± 118.40 | < **0.001** | 0.080 | 723.00 ± 133.13 | 677.56 ± 121.80 | **0.022** |
|  | Reaction times in incongruent stimuli | 751.79 ± 148.77 | 663.61 ± 149.02 | < **0.001** | 0.090 | 764.28 ± 152.17 | 721.14 ± 140.05 | **0.049** |
|  | Stroop interference effect | −23.01 ± 73.67 | −7.09 ± 44.24 | 0.217 | 0.010 | 3.20 ± 145.73 | −8.53 ± 63.52 | 0.616 |
| ***Response inhibition***<br>*(Go/No-Go task)* | Accuracy in Go trials | 0.83 ± 0.14 | 0.94 ± 0.08 | < **0.001** | 0.120 | 0.82 ± 0.15 | 0.82 ± 0.15 | 0.968 |
|  | Reaction times in Go trials | 486.47 ± 70.21 | 426.96 ± 49.87 | < **0.001** | 0.160 | 489.80 ± 71.90 | 498.17 ± 74.78 | 0.507 |
|  | Accuracy in No-Go trials | 0.83 ± 0.12 | 0.88 ± 0.11 | 0.166 | 0.010 | 0.84 ± 0.13 | 0.84 ± 0.14 | 0.840 |
| **Dimension II: Working memory updating and maintenance** |  |  |  |  |  |  |  |  |
| ***Updating***<br>*(Running memory task)* | Accuracy in 1,750 ms | 0.60 ± 0.25 | 0.74 ± 0.21 | < **0.001** | 0.060 | 0.56 ± 0.26 | 0.52 ± 0.28 | 0.395 |
|  | Accuracy in 750 ms | 0.51 ± 0.29 | 0.66 ± 0.22 | **0.001** | 0.050 | 0.49 ± 0.29 | 0.44 ± 0.27 | 0.258 |
| ***Maintenance***<br>*(Digit span backward; Corsi block test)* | Span in digit span backward task | 5.07 ± 1.51 | 6.06 ± 2.11 | < **0.001** | 0.090 | 5.05 ± 1.65 | 5.31 ± 1.76 | 0.308 |
|  | Span in Corsi block test | 4.50 ± 1.00 | 4.76 ± 1.27 | 0.101 | 0.010 | 4.56 ± 0.99 | 4.84 ± 1.13 | 0.258 |
|  | Accuracy in Corsi block test | 0.53 ± 0.20 | 0.55 ± 0.21 | 0.480 | 0.000 | 0.54 ± 0.21 | 0.56 ± 0.20 | 0.616 |
| **Dimension III: Cognitive flexibility/Switching** |  |  |  |  |  |  |  |  |
| **Switching**<br>(Number switching task) | Switching cost | 304.80 ± 303.40 | 198.95 ± 238.37 | **0.001** | 0.030 | 292.36 ± 300.00 | 277.50 ± 286.88 | 0.840 |
|  | Mixing cost | 62.43 ± 189.87 | 112.30 ± 149.69 | 0.278 | 0.010 | 70.90 ± 213.90 | 14.32 ± 213.43 | 0.258 |
| **Composite scores** |  |  |  |  |  |  |  |  |
|  | Inhibition composite | −11.09 ± 36.83 | −3.10 ± 22.12 | 0.217 | 0.010 | 2.02 ± 72.87 | −3.85 ± 31.75 | 0.484 |
|  | Executive function composite | 98.10 ± 102.30 | 65.53 ± 80.06 | **0.001** | 0.030 | 98.31 ± 103.95 | 91.39 ± 94.78 | 0.624 |
|  | Inhibition and updating composite | −5.24 ± 18.43 | −1.18 ± 11.05 | 0.217 | 0.010 | 1.29 ± 36.43 | −1.67 ± 15.89 | 0.481 |
|  | Inhibition and switching composite | 146.86 ± 153.44 | 97.92 ± 120.07 | **0.001** | 0.030 | 147.19 ± 155.91 | 136.82 ± 142.15 | 0.625 |
|  | Updating and switching composite | 152.70 ± 151.71 | 99.84 ± 119.20 | **0.001** | 0.030 | 1146.46 ± 150.01 | 139.01 ± 143.46 | 0.729 |

**Note:** The $p$ -values in bold face indicate statistically significant differences ( $p < 0.05$ ). The $p$ -values were adjusted using the false discovery rate (FDR) with Benjamini–Hochberg procedure. $\eta^2$ , eta-squared effect size. The interference effect is quantified as the difference in reaction time between incongruent and congruent trials in the *Stroop* task.

Symptom severity in each patient with schizophrenia was evaluated using the PANSS^87^. Scores for four symptom dimensions (i.e., positive, negative, affective, and cognitive factors) were derived for each patient via the Dimensions and Clustering Tool for Schizophrenia Symptomatology (DCTS; http://webtools.inm7.de/sczDCTS/). These dimensions have been previously identified as stable and generalizable across populations, regions, and clinical settings^20^. Higher scores denote more severe symptoms within each dimension. Patients were further categorized into the positive subtype, negative subtype, or ambiguous cases lying in-between these two subtypes based on their DCTS-derived symptom dimensional scores and membership values. We employed a heuristic membership degree of 0.6 as the cutoff value for the ambiguous cases subgroup given that these participants were not clearly assigned to any of the two more differentiated negative-positive subtypes.

For the 86 followed up patients with schizophrenia, remission versus non-remission prognostic statuses were determined based on the RSWG criteria^34^. Remission is defined by scores ≤ 3 on key PANSS items (P1, P2, P3, N1, N4, N6, G5, G9). Considering the current lack of consensus on a definition of clinical outcomes in schizophrenia, we utilized an alternative definition. This definition was based on the reduction of the total PANSS score, and three remitted or non-remitted conditions were specified by: 1) a 25% reduction; 2) a 35% reduction; and 3) a 50% reduction^36^. Clinical outcomes of patients were moreover defined based on subtype-membership transition: 1) baseline non-negative subtype with transition to a negative subtype, and 2) stable negative subtype during follow-up. This definition helps in identifying patients that experience primary (and stable) negative symptoms or secondary symptoms due to antipsychotic treatments and related factors^38,39^.

### Assessments

#### Sociodemographic and electronic records

The standard 60-item Raven’s Progressive Matrices test was used to assess fluid intelligence^88^. Family socioeconomic status was assessed using a family financial status questionnaire^89^. Thereafter, we collected detailed clinical information from the electronic medical records of patients (e.g., disorder onset, number of episodes, age at diagnosis, duration of illness, types and dosages of antipsychotic drugs, family medical history).

#### EF

This study was based on the influential model subdividing EF into three core dimensions: inhibitory control; working memory (updating and maintenance); and cognitive flexibility/shifting^15,90^. Working memory updating is the process of continuously replacing old information with new in working memory, according to current task requirements. Working memory span/maintenance is the ability to maintain and process information over a period of time, often directly linked to short-term memory capacity^40^. Inhibitory control involves the ability to suppress dominant responses and adapt to a changing environment, minimizing the impact of irrelevant information on ongoing information processing^15^. Therefore, inhibition is also divided into two dimensions: interference inhibition (or interference control) and response inhibition (or behavioral inhibition)^91^. Cognitive flexibility is considered a single dimension, characterizing the ability to flexibly switch between different tasks and modes of thought^92^.

According to their complexity, we selected six behavioral tasks to measure these EF dimensions (Fig. 3)^40^: 1) *number running memory updating* task was used to examine working memory updating^89^; 2) *digit span backward* task was used to measure working memory maintenance (span)^40^; 3) *Corsi block* test was used to measure working memory maintenance (span), which more comprehensively assesses maintenance in the spatial dimension^93^; 4) *Stroop* task was used to measure interference inhibition^94^; 5) *Go/No-Go* task was used to measure response inhibition^95^; and 6) *number switching* task was used to measure shifting^92^. All behavioral tasks were performed using *E*-Prime 3.0 software (Psychology Software Tools, Inc., Pittsburgh, PA, USA). Accuracy and reaction time on each task can be weighted to derive 14 comprehensive assessment indicators from these six behavioral tests.

#### Inhibitory control

The four measurements for assessing the interference control function included the reaction times for the incongruent, congruent, and neutral stimuli, and the difference of reaction time between the congruent and the incongruent condition trials (i.e., the interference effect) in the *Stroop* task. Three measurements for assessing the response inhibition function included the reaction time for the “Go” stimuli and the accuracy for the Go and No-Go stimuli in the *Go/No-Go* task.

#### Working memory

Two measurements for assessing working memory updating function included the proportion of digits correctly recalled and placed in the correct sequence at two different speeds of presentation (1,750 ms and 750 ms per digit) in the *running memory* task. Three measurements for assessing the numeric working memory maintenance capacity included the length of the last correctly repeated sequence, the count of sequences correctly repeated until the conclusion of the test (i.e., the total number of successful trials) from the *Corsi block* test, and the maximal number of digits accurately recalled in the reverse order of the *digit span backward* task.

#### Cognitive flexibility

Two measurements which included the difference in reaction time between the switch and the non-switch trials [switch cost], as well as the difference in the reaction time between the non-switch and the single-task trials [mixing cost]) measured in the *number-letter switching* task.

Besides a conceptual formulation of the 14 task measurements according to the three-dimensional representation of EF, we supplemented five composite scores calculated based on these measured variables^96^. Firstly, an inhibitory composite score was calculated by averaging: 1) the difference in reaction time between the congruent and the incongruent condition trials in the *Stroop* task; and 2) the accuracy for the No-Go stimuli in the *Go/No-Go* task as in previous studies^97,98^. The purpose of this approach was to denote the combined response and inference inhibitory functions. Furthermore, this inhibitory composite score was aggregated with the assessments in the *running memory* task (the proportion of digits correctly recalled and placed in the correct sequence at the speed of 1,750 ms per digit) and the *number-letter switching* task (switch cost) to form an abbreviated version for representing general EF functions. Such abbreviation is in compliance with the previous notion on a single-condition indicator that these trails require greater executive control demands^99^. Considering that EF functions interplay across conceptual constructs during cognitive engagement (e.g., problem-solving) for processing particular behaviors^100^, we additionally created three cross-dimensional EF composite scores by collapsing the cardinal three dimensions of EF, which the inhibitory composite score was similarly used: 1) Inhibition and Updating composite; 2) Inhibition and Switching composite; and 3) Switching and Updating composite. The measurements used to assess working memory updating and switching were employed to calculate the abbreviated version of the EF composite score. These composite measures would provide additional insights into the dimensional and cross-dimensional contributing features to diagnostic and prognostic clssifications^100^. Consequently, 19 EF-related indicators were used as input features for the machine learning models (Fig. 3).

### Behavioral task and clinical scale analyses

We performed a mixed-model ANCOVA with group (schizophrenia vs. healthy control) as a between-subjects factor and all EF measurements as well as composite scores as within-subjects factors, while controlling for sociodemographic variables, to examine whether schizophrenia has differentially affected EF performance. The mixed-model ANCOVA was followed by multiple one-way ANCOVAs to examine group differences in each EF measure. also controlling for Sociodemographic variables were likewise controlled and a Benjamini-Hochberg (BH) false discovery rate (FDR) correction was applied to adjust for multiple comparisons. Eta squared effect sizes were calculated to describe the magnitude of effect sizes, with certain values interpreted as small (*η²* ≤ 0.01), medium (0.01 < *η²* ≤ 0.06), or large (0.06 < *η²* ≤ 0.14)^101^. Two-sample t tests were used to compare clinical symptoms, demographic characteristics, and EF between the remission and non-remission subgroups. Chi-squared tests were used to compare categorical variables between these groups. We used paired-samples *t*-tests to compare clinical symptoms between baseline and follow-up (after 4–6 weeks of treatment) for the subset of followed participants. Finally, we used Pearson and Spearman correlation analyses to examine the relationships among clinical symptoms and EF, with the former applied to continuous variables and the latter applied to categorical variables.

### Classification modeling procedure

#### Features and models

Participants were categorized into schizophrenia and HC groups. Two feature sets were tested for diagnostic classification accuracy: 1) 19 baseline EF assessments, subsuming the three EF dimensions measured by six behavioral paradigms (Table 2); and 2) 32 features—the 19 baseline EF measures plus 13 routinely attainable sociodemographic variables. For prognostic classification, the same two feature sets were tested to distinguish patient remission status (remitted vs. non-remitted) after 4–6 weeks of antipsychotic treatment. SVM, RF, and AdaBoost, which are widely used in psychiatric machine-learning research^102^, were used for classification tasks, along with a synthesized stacking model of the three. Stacking models are a multi-view approach integrating classification weight estimates from single classifiers to improve ultimate performance^103^.

### Complementary investigations

#### Control analysis

Several control analyses were performed, in which we controlled the effects of 1) 13 demographic characteristics; 2) both the 13 demographic variables and the OZP equivalent dosage; 3) only the OZP equivalent dosage on the baseline EF assessments using regression approaches^104^ when establishing the diagnostic and prognostic classification models. The continuous variables in the demographic dataset included age, years of education, body mass index, socioeconomic status, and fluid intelligence measured by Raven’s Progressive Matrices test. The categorical variables consisted of sex, ethnicity, residence, employment status, only-child status, marital status, smoking history, and drinking history. One-hot encoding was applied to the categorical variables, converting them into binary vectors. Furthermore, we sought to mitigate the potential impact of antipsychotic drug dosage and evaluate the robustness of our classification results. Thus, we established the prognostic classification models with only those patients receiving a clinically standard, commonly effective OZP equivalent dosage of 10–20 mg/day.

#### Sensitivity analysis

In our research, the majority of dropouts were due to hospital discharge. Therefore, we conducted a best-case sensitivity analysis, assuming that all participants who were lost to follow-up had positive treatment outcomes (remission). Additionally, we performed a worst-case analysis, assuming that all participants who were lost to follow-up had the least favorable treatment outcomes (no remission). These data help establish the potentially extreme scenarios due to attrition with respect to our main analyses with an observed prognostic status. Sensitivity analyses, such as best-worst (assuming all participants lost to follow-up in one group [referred as group 1] have had a beneficial outcome and all those with missing outcomes in the other group [group 2] have had a harmful outcome) and worst-best case (assuming that all participants lost to follow-up in group 1 have had a harmful outcome; and that all those lost to follow-up in group 2 have had a beneficial outcome), are utilized in medical and psychiatric research to evaluate the reliability of results in relation to participants who do not complete the study^105,106^.

### Machine learning design

Machine learning and CV were implemented using Python (version 3.10.11) and the scikit-learn package (version 1.3.0). Specifically, the original data were first preprocessed to accommodate missing values, outliers, and class imbalance issues (see Supplementary Materials for details). Thereafter, we randomly split the preprocessed data into a discovery dataset with 80% of the overall schizophrenia and HC groups (‘training set’) and a ‘lock-box’ test dataset with the remaining 20% of these samples (‘test set’) to determine out-of-sample classification performance(Fig. 1)^4,42–44,107^. The random split procedure was stratified for the outcome variable (diagnostic label or remission status), ensuring a balanced representation of labels in each dataset^42,43^. Using the discovery dataset, we performed a nested CV loop^108^ (also termed double CV), which differentiates two CV roles to avoid ‘circularity’ introduced by overfitting when the same sample subset is used for both hyperparameter tuning and model validation^53^. Specifically, within a nested CV loop, the inner CV (k= 3), encompassing 80% of the discovery sample, operates all data-dependent decisions while determining optimal hyperparameters. The outer CV (k= 5) is subsequently utilized for parameter assessment and model selection^78^. For optimal hyperparameter selection, we used the Optuna optimization technique (version 3.5.0) and selection based on the achievable AUC of candidate hyperparameters within the validation sets of the inner loop^109^. The AUC metric, representing the degree of separability, is widely used to evaluate model performance. In this investigation, it indicated the ability of the model to distinguish between the schizophrenia and HC groups, and between the patient remission and non-remission groups. Hyperparameters with the highest average performance over the 5 × 3 nested CV were used to train a model on the entire discovery sample without further modification; next, they were tested using the independent ‘test set’ sample^50^. In addition to AUC, sensitivity, specificity, and balanced accuracy performance metrics were assessed^110^. To avoid potential bias from random splitting, the aforementioned machine learning procedure was repeated 100 times^111,112^.

#### Feature importance analysis

To evaluate the contributions of EF features to our classification models, we assigned an importance score (i.e., Shapley value) to each feature^47^. Specifically, we used the SHAP library (version 0.39.0) model-agnostic SHAP KernelExplainer approach, which is generally used to estimate Shapley values for prediction models^113^. SHAP KernelExplainer employs a Monte Carlo approach to randomly sample feature combinations based on input predictors. Initially, it estimates the importance of these combinations with varying features present in model predictions. Subsequently, individual Shapley values are calculated to denote the contribution of each feature to the target prediction based on a weighted linear regression model^48^.

After obtaining the individual-level Shapley values for each feature, we computed the mean absolute Shapley value (i.e., feature importance score) across all individuals (i.e., the group-level Shapley values) where larger Shapley values indicate stronger importance of this feature to the classification model. The group-level Shapley values were next used to depict a group-level decision path for each feature. Essentially, among those participants in the schizophrenia group who were correctly classified, performance by each EF dimension assessment was averaged across both the HC and schizophrenia groups for the diagnostic models, and across the remission and non-remission patient groups for the prognostic models. The averaged performance of each EF feature was then normalized and compared with a positive (or negative) value; higher (or lower) performance by an EF feature drove the model to more accurately classify true cases. Using individual Shapley values, we also plotted the decision path for each individual who was correctly classified to complement the group-level results; this was performed because individuals with schizophrenia have heterogeneous expressions across EF dimensions^60^. Finally, based on the individual Shapley values, we tested the extent to which the importance of the contribution of an EF feature to a classification was linked to individual psychopathology. This was conducted using Pearson correlation analysis based on individual scores (entire patient group *N*=195, diagnostic model; subset sample *N*= 86, prognostic model) along the four symptom dimensions which were assessed at baseline and follow-up, and the difference between the two. The FDR approach was used for the correction of multiple comparisons to statistically rule out potential false-positive associations^114^.

## Acknowledgements

This work was supported by the National Natural Science Foundation of China (No. 32260207 [to Xin Zhao], No. 82371506 [to Ji Chen] & No. 82201658 [to Ji Chen]), the STI2030-Major Projects (No. 2022ZD0214000 [to Ji Chen]), and the National Key R&D Program of China (No. 2021YFC2502200 [to Ji Chen]). BTTY is supported by the NUS Yong Loo Lin School of Medicine (NUHSRO/2020/124/TMR/LOA), the Singapore National Medical Research Council (NMRC) LCG (OFLCG19May-0035), NMRC CTG-IIT (CTGIIT23jan-0001), NMRC STaR (STaR20nov-0003), Singapore Ministry of Health (MOH) Centre Grant (CG21APR1009), the Temasek Foundation (TF2223-IMH-01), and the United States National Institutes of Health (R01MH120080 & R01MH133334). Any opinions, findings and conclusions or recommendations expressed in this material are those of the authors and do not reflect the views of the Singapore NMRC, MOH or Temasek Foundation.

## Conflict of interest

The authors declare no conflicts of interest.

## Data availability statement

Information for the main sample used in the present study have been included in the Supplementary Materials. The raw data of our used sample are protected and are not publicly available due to data privacy. These data can be accessed upon reasonable request to the corresponding author (X. Z.). Derived data supporting the findings of this study are available from the corresponding authors (X. Z. or J. C.) upon request.

## Code availability statement

Scripts to run the main analyses have been made publicly available and can be accessed at https://doi.org/10.6084/m9.figshare.26086594.v1.

## Notes

### Competing Interest Statement

The authors have declared no competing interest.

### Author Declarations

The Ethics Committee of the Third People's Hospital of Lanzhou City and the Ethics Committee of Northwest Normal University (Lanzhou, China) gave ethical approval for this work. This study adhered to the principles outlined in the Declaration of Helsinki. All participants provided written informed consent prior to participation.

